# The landscape of polygenic score prediction accuracy and transportability across the phenome in ancestrally diverse populations

**DOI:** 10.64898/2026.10.08.26365087

**Authors:** Meng Lin, Christopher H. Arehart, Simon N.K. Walker, Nicholas Rafaels, Kristy R. Crooks, Matthew Fisher, Sameer Chavan, Harry Smith, Nikita Pozdeyev, Leslie A. Lange, Milton Pividori, Jonathan A. Shortt, Ryan M. Layer, Colorado Center for Personalized Medicine, Audrey E. Hendricks, Sridharan Raghavan, Christopher R. Gignoux

**Affiliations:** Department of Biomedical Informatics, School of Medicine, University of Colorado Anschutz; Colorado Center for Personalized Medicine, University of Colorado Anschutz; Institute for Behavioral Genetics, University of Colorado Boulder; Department of Ecology & Evolutionary Biology, University of Colorado Boulder; BioFrontiers Institute, University of Colorado Boulder; Department of Computer Science, University of Colorado Boulder; Human Medical Genetics and Genomics Graduate Program, University of Colorado Anschutz; US Department of Veterans Affairs Eastern Colorado Health Care System

## Abstract

Polygenic scores (PGS) are increasingly common in human genetics research, yet their performance and portability vary substantially across traits and populations. We systematically evaluate prediction accuracy and cross-ancestry portability for 3,994 published PGSs across the phenome in the ancestrally diverse CCPM biobank. We identify widespread trait-specific and cross-trait associations, demonstrating both the utility and breadth of existing polygenic predictors. However, predictive effects frequently differ across ancestry groups, highlighting persistent challenges for portability that threaten equity in downstream applications. We further find that accuracy and portability depend strongly on whether the PGS is continuous or thresholded. Finally, through hierarchical mixed modeling, we identify score-development, genome features, and cohort characteristics associated with improved PGS accuracy and portability. These results highlight key factors supporting future development of robust and portable scores for research and clinical translation.

## Introduction

Polygenic scores (PGSs) have come to the forefront of human genetics as an efficient way to summarize and condense genetic risk for complex traits. PGS-based prediction is used to address research questions in evolution(Berg and Coop 2014; Novembre and Barton 2018; Berg et al. 2019), epidemiology (Hujoel et al. 2022; Darst et al. 2021; Mars et al. 2020), and personalized medicine ^4,5^. Predicting traits by leveraging PGS alone or in conjunction with other information can be highly significant, approaching or exceeding prediction ability from family-based measures as sample sizes become very large^1^. Accordingly, PGS are attractive as tool to identify individuals with elevated risk of a clinical condition, followed by standard clinical screening, facilitating earlier diagnosis and improved disease management, for example in settings where subtype misclassification is common such as distinguishing adult-onset type 1 diabetes (T1D) from type 2 (T2D) ^6–8^. For some major clinical endpoints, PGSs have been shown to have equal or even greater potential for identifying at-risk individuals than monogenic testing ^2,3^.

Despite promising utility, inconsistent portability of PGS across genetic ancestries continues to be a major impediment to broader clinical and translational implementation. Like most prediction models, PGSs have diminished prediction accuracy in populations that differ from the populations in which they were derived ^9,10^. Additionally, population structure can confound results ^11^. These portability challenges are considered to have multiple sources, including but not limited to linkage and allele frequency patterns and effect size heterogeneity due to GxG or GxE interactive components (Martin et al. 2017, 2019; Wojcik et al. 2019; Bitarello and Mathieson 2020; Kanai et al. 2021; Mathieson 2021; Hou et al. 2023; Hu et al. 2025).

Given these challenges and burgeoning potential applications, a natural question is to assess the quality of PGS performance in real-world settings. Previous approaches have compared performance among candidate PGS methods and varying demographic compositions of cohort populations, typically focused on the traits for which a PGS was developed (e.g., type 2 diabetes PGS to predict type 2 diabetes). ^12–19^. Complementary to such “on-target” comparisons, some health outcomes may benefit from PGS developed for genetically related traits (i.e., “off-target” approaches) ^20–22^. This may be particularly useful in situations when “on-target” PGS of a trait of interest is limited, whereas PGSs of traits that are genetically correlated to the target, sharing etiology, or residing in the same causal path to an outcome are readily available. However, systematic benchmarking of off-target PGS associations is still underexplored, despite the promising translational and clinical potential that would be missed by ignoring the spectrum of genetic correlation among complex traits^23^.

Amid continued development of new PGS models, more practical and perhaps fundamental questions emerge as well: what are features underlying traits, study designs, or score methodologies that predict a better or worse PGS performance in terms of accuracy and portability? Answers to this question can offer scientific insight about genetic prediction and guidance for future PGS development.

Driven by these combined motivations, we relied on the wealth of existing scores available in the PGS Catalog ^24^ and evaluated associations of > 5.7 million PGS-trait pairs across the phenome of >1,400 electronic health record (EHR) diagnoses in over 73,000 individuals spanning six genetic similarity groups that reflect continental ancestry origins from the biobank at the Colorado Center for Personalized Medicine (CCPM)^25^. We then used a data-driven and trait-agnostic analysis to identify the attributes of scores, traits, and cohorts that lead to high accuracy and portability of PGS performance. We demonstrate the limitation of poor-performing scores in multi-trait analyses such as Mendelian Randomization. These insights offer guidance for using current scores and for developing the next generation of PGS with improved clinical and translational potential for all.

## Results

### Overview of phenotype-PGS associations

The recently established biobank at CCPM, a large institutional biobank co-developed by the University of Colorado and its partner health system, UCHealth, represents an out-of-sample dataset to evaluate previously published PGS. CCPM has catchment across the United States with enrichment in the Mountain West region and includes individuals from ancestrally diverse backgrounds. We retrieved all 4,742 PGS scoring files available from the PGS catalog ^24^ (https://www.pgscatalog.org, accessed in May 2024) and applied them to 73,346 (69,515 unrelated) individuals in the CCPM biobank freeze3 ^25,26^ using our PGS harmonization and calculation pipeline (Methods). This resulted in 3,994 scores developed across 1,311 reported traits, or 584 Experimental Factor Ontology (EFO)-labeled traits being successfully calculated in CCPM samples (Supplementary Table 1).

We completed phenome-wide-by-PGS associations within each of the six genetically defined similarity groups in freeze3 ^26^. This allowed us to compare performance across genetic ancestry groups rather than adjusting out the effects of population structure ^27^ (and see Discussion). The genetic similarity groups (GSGs) in CCPM were named after their most closely matched reference group from the 1000 Genomes (TGP) and Human Genome Diversity Project (HGDP), reflecting broad continental ancestry related to Africa (AFR), Hispanic/Latino or America (AMR), Central South Asia (CSA), East Asia (EAS), Europe (EUR), and Middle East (MLE). GSGs in CCPM thus include TGP/HGDP-AFR-like (unrelated N=2,859), TGP/HGDP-AMR-like (unrelated N=8,126), TGP/HGDP-CSA-like (unrelated N=1,162), TGP/HGDP-EAS-like (unrelated N=1,401), TGP/HGDP-EUR-like (unrelated N=55,721), and TGP/HGDP-MLE-like (unrelated N=246). We evaluated associations using two transformations of PGS: 1) standard normalization (named “*continuous*” in the following context) and, 2) stratified at the top decile compared to the remaining 90^th^ percentile (named “*thresholded*” in the following context). All associations accounted for common covariates of age, sex, genotyping batch, and the top 10 genetic principal components (PCs). This was followed by meta-analysis (Methods) across GSGs, resulting in over 70M association pairs. To avoid potential collinearity of PGSs with genetic relatedness, we filtered to a maximal set of unrelated individuals and employed Firth regression to account for case-control imbalance (Methods). Associations passing a Benjamini-Hochberg FDR<0.1 are defined as our *significant set* (n = 371,196 and 206,842 for *continuous* and *thresholded* PGS-trait pairs, respectively). Unless otherwise defined, we used these significant sets (with a focus on the *continuous*, most common use-case) for downstream analyses. Below, we report evaluations of PGS prediction accuracy and portability based on trait-score associations per GSG and our combined population meta-analyses. An overview of the study design is presented in Figure 1a.

**Figure 1.**
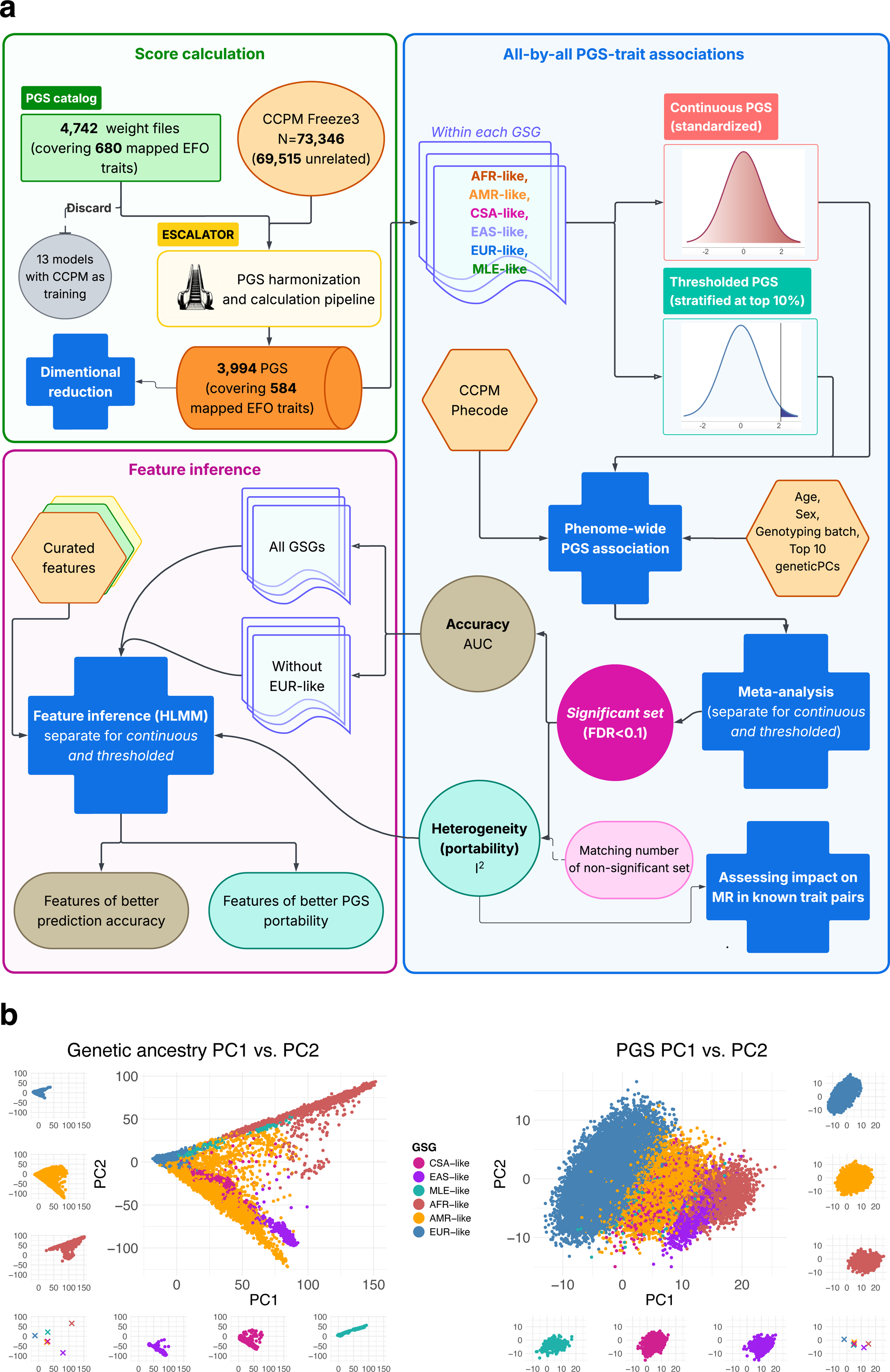
PGS models are applied and evaluated across CCPM biobank participants. **a**, A scheme overview of major designs and analyses in this study. **b**, PCA of genetic data (left) and PCA of PGSs (right) in CCPM biobank participants. (Left) PCA of the Freeze3 genetic dataset of CCPM biobank, illustrating major population structures. Colors and subpanels are constructed the same way as in the left to mirror the corresponding information. (Right) After pruning out redundant PGSs designed for the same phenotype, PCA was performed on 518 “independent” scores over CCPM participants. The first two PCs are shown in the main panel with participants colored by ancestry-informed similarity groups they are assigned to. Sub panels on the left and bottom separately show each group’s individuals for an unblocked view. Sub panel on the lower left shows the geometric mean averaged within each group.

### Variance among PGSs is largely explained by between genetic similarity group differences and within-EUR-like substructure

We summarized the overall pattern of the PGSs by applying principal component analysis (PCA) after pruning for the most significant PGS per trait down to 541 scores to avoid bias due to over-or under-representation of scores across phenotype domains. We found the primary PGS-PCs recapitulated genetic ancestry as observed with genetic PCA (Figure 1b): PGS-PC1 particularly showed a distinct gradient between EUR and AFR genetic ancestries, clearly mirroring genetic-PC1; PGS-PC2, in contrast to genetic-PC2, reflects substructure in EUR groups (Supplementary Table 2). Over 80% (3,355 over 3,994) of PGSs use either GWAS summary statistics or a training cohort of only European descendants. Thus, PGS-PC2 primarily capturing within-EUR-like variation likely reflects the over-representation of European genetic ancestry in the creation of PGSs. Despite this difference in PC2, the top 5 PGS and genetic PCs were strongly and significantly correlated (e.g. Pearson’s correlation coefficient r=0.85 for PC1, p<4.94e-324, Supplementary Table 3).

### Characterizing phenome-wide PGS associations and visualization of top association results

The PGSs were originally developed for 584 unique phenotype strings spanning 17 phenotypic categories as annotated in the PGS Catalog (including one extra “unassigned” category containing 46 ontology identifiers). PGSs were unevenly distributed across these categories.

Traits relating to “Cancer” and “Lipid or lipoprotein measurement” are particularly common (Figure 2). In testing outcomes in CCPM, we used phecode mapping (https://phewascatalog.org/phewas/) ^28,29^ to generate 1,439 codes with case counts >10 across 18 phecode domains in CCPM (the unassigned category containing 14 phecodes). Overall, all 3,994 PGSs were significantly associated with at least one phecode when using *continuous* PGS models (at FDR<0.1, or p< 6.46e-3), and 3,989 out of 3,994 (99.9%) when using *thresholded* PGS models (at FDR<0.1, or p<3.60e-3).

**Figure 2.**
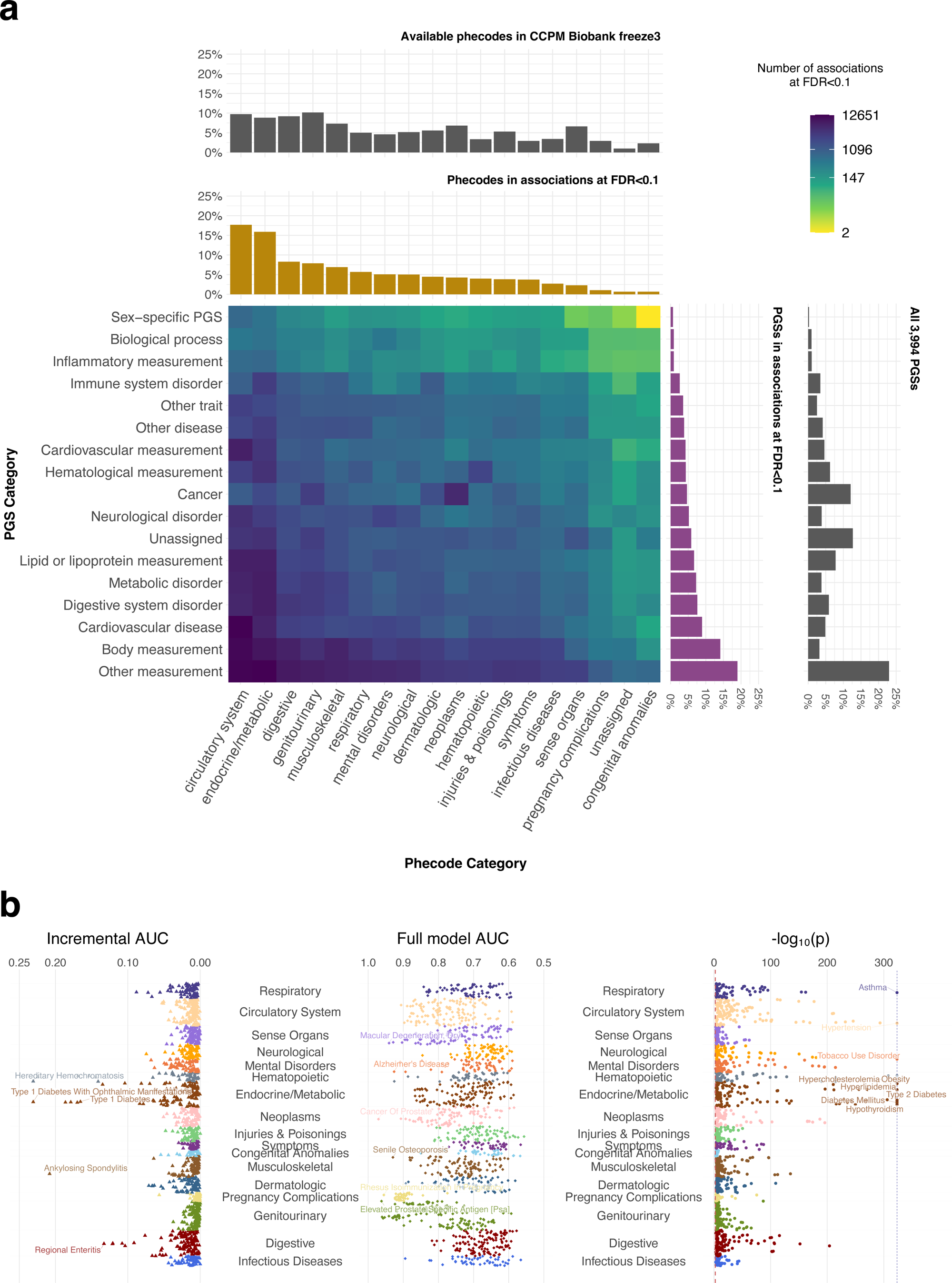
Significant PGS by phecode associations populated common disease domains. **a**, Phecode, PGS, and phecode by PGS distributions across phenotype domains before and after (FDR<0.1) associations. Phecodes derived from ICDs distribute unevenly across 17 clinical domains (grey bars on the top, with domain names at the bottom), and 3,994 PGSs adopted from the PGS catalog spread over 15 trait categories (grey bars on the right, with category names on the left). Among 10,935 significant associations (FDR<0.1, PGS taken as continuous unit), the distribution of phecode and PGSs shifted (yellow and purple, respectively). The heatmap denotes counts of significant associations falling into each phecode domain by PGS category pair. **b**, Manhattan plots of phecode ∼ PGS association signals, shown in incremental AUC by PGS (left), full model AUC (middle), and log-transformed P-values (right). PGS is taken as the quantitative measure, and association models correct age, sex, genotyping batch, and top 10 PCs. Each datapoint represents a single phecode with its most significant PGS. The lowest p-values on the right only is the smallest double float value.

Focusing on the *continuous* PGS models, the phecode categories with the largest number of significant associations (out of 371,196 total PGS x phecode pairs in the *significant set*) were “circulatory system” and “endocrine/metabolic” (66,164 [17.8%] and 59,126 [15.9%] respectively, Figure 2). Over 80% of the strongest associations (defined as p<1e-100) appeared in these two phecode domains (encompassing 51 phecodes, Supplementary Table 4). The top assigned PGS categories were “Body measurement” (e.g., body mass index and waist-hip ratio), and “Digestive system disorder” (Figure 2a, Supplementary Table 5).

As expected, we observed that phecode domains frequently had significant predictions from PGSs designed for the same phenotypic category. For example, phecodes under “neoplasm” were more frequently associated with PGSs designed for “cancer”, “hematopoietic” phecodes with PGSs for “hematological measurement”, “endocrine/metabolic” phecodes with PGSs for “metabolic disorder”, and “circulatory system” phecodes with PGSs for “cardiovascular disease” (Figure 2a).

Most strong PGS-phecode associations occurred with diagnoses in the “endocrine/metabolic” category, including but not limited to T2D, obesity, and hypercholesteremia (all p < 4.94e-324); outside this category, traits including hypertension (HT), tobacco use disorder, and asthma exhibited a similar level of significance in associations (Figure 2b; right panel). We note that, in general, these exceptionally small p-values result from a combination of common diseases in CCPM (e.g. prevalence of 40%, 16%, and 35% for HT, asthma, and tobacco use disorder, respectively), relatively high trait heritability, and higher predictive accuracy of the PGS.

We also evaluated model prediction performance using area under the curve (AUC). When incorporating additional covariates such as sex and age, the top full model associations were readily found among symptoms with large age-related influences, such as macular degeneration, Alzheimer’s disease, prostate cancer, and senile osteoporosis (Figure 2b; middle panel). To look at the relevant effects of PGS directly, we measured the incremental AUC (Methods). Top signals of incremental AUC arose in traits characterized by large genetic effects, such as hereditary hemochromatosis, T1D (and related sequelae), ankylosing spondylitis, and regional enteritis/Crohn’s disease (Figure 2b; left panel). The full series of top hits and associations with further details are presented in the summary table and its PGS-trait visualizer described in the Data Availability Statement.

We highlight a preliminary list of candidate PGSs that showed promising performance (p<1e-50 using either *continuous* or *thresholded* PGS units, with incremental AUC > 5%) and low heterogeneity (I^2^=0) across the GSGs in CCPM (Supplementary Table 6). The candidate PGSs are potentially worth consideration for translational utility given their balanced accuracy and portability. The majority identified by the list appeared to be autoimmunity-related outcomes, including T1D and related symptoms, ankylosing spondylitis, and Celiac disease, while the top associated phenotypes identified merely by very small p-values did not emerge based on this combination of selection criteria.

We confirmed PGSs’ potential to identify individuals at elevated risk for common diseases ^3^ using five well-studied, prevalent health outcomes as key examples: prostate cancer, female breast cancer, T2D, HT, and obesity. The most-significantly associated PGS for each of the five conditions was able to classify almost half (42.7%-49.8%) of the CCPM participants into a high-risk category under the corresponding condition, characterized by having at least two-fold increase in the odds of disease development (i.e. OR>2) relative to the lower-risk remainder of the biobank, with estimates adjusted for demographic covariates. Further investigation of the high-risk tail of the distribution yielded stronger effects: about 9% of total male participants had OR>5 of elevated risk in prostate cancer than the rest of participants, while 11% of total female participants had OR>3 of breast cancer. Other domains showed attenuated effects, though the threshold of OR>3 still yielded more than 5% of CCPM participants in the high-risk group both for T2D and HT (Supplementary Table 7).

Highlighting the top association results, we provided an interactive web tool visualizing the per-GSG and meta-analyzed summary statistics inquired by either PGS or phecode, aside from a full release of all by all summary statistics in this study (see Data Availability Statement).

### Heterogeneity of PGS performance across GSGs and its impact in causal relationship inference

We next assessed PGS portability across GSGs in CCPM based on effect size meta-analysis heterogeneity (quantified as the I^2^ statistic ranging from 0 [low] to 1 [high]) and impact on downstream applications of PGS, such as Mendelian randomization (MR) (see Supplementary Notes for choices of heterogeneity estimates).

Among the *significant set*, we observed an average I^2^ = 0.22 (IQR: 0.00-0.43) for *continuous* PGS units (n=371,196) and 0.20 (IQR: 0.00 – 0.40) for *thresholded* (n=206,842) (Supplementary Figure 2). We found low concordance of I^2^ for *continuous* and *thresholded* transformation (Lin’s Concordance Correlation Coefficient =0.369 for 154,043 associations passing FDR<0.1in both units, Figure 3a, Supplementary Figure 3), suggesting that portability is likely affected by choice of PGS transformation.

**Figure 3.**
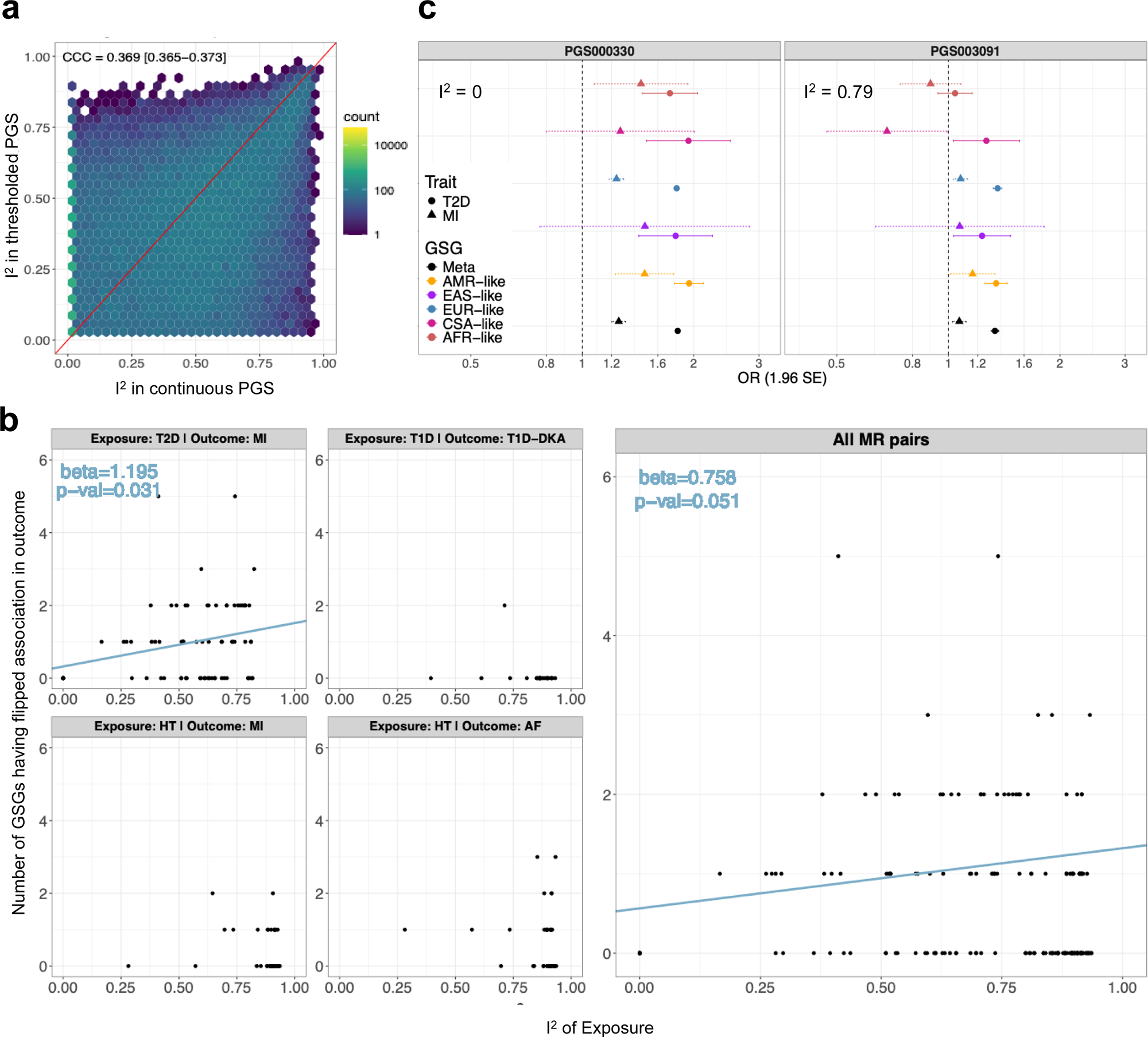
Heterogeneity of effect size across GSGs varies and impacts inference of causal relationship of traits. **a**, Correlation of I^2^ in PGS-trait pairs passing FDR<0.1 in both continuous (x-axis) and thresholded (y-axis) units. **b**, Number of GSGs in the secondary outcome exhibiting unexpected flip-flops in PGS association effect (y-axis), against I^2^ of PGS on the exposure trait (x-axis), within four selected MR pairs (left sub panels), and all pooled pairs (right large panel). Regression line and coefficients are shown in blue if the regression is at least borderline significant. **c**,Two scores with different heterogeneity performances on T2D (I^2^ denoted in each sub panel) were associated with T2D (circle shaped) and its downstream complication of MI (triangle shaped). Effect size is present in OR, with error bars representing 95% C.I.

A common research application of PGS is assessing causal relationships among traits through MR, where a PGS is used as a genetic instrumental variable (IV) ^30–32^. We hypothesized that PGSs with higher heterogeneity of effect sizes across GSGs is more likely to be confounded by population structure and would propagate that variability of effect sizes through MR estimates, inducing erroneous causal inference. We tested this by examining consistency of direction of effect estimated in two-stage least-squares MR analyses of four exemplary trait pairs with established causal relationships *and* all candidate PGS IVs developed for the corresponding exposure trait within each pair: (1) T2D as a risk factor for myocardial infarction (MI) ^33^, (2) T1D as a risk factor for T1D with ketoacidosis (T1D-DKA)^34^, (3) HT as a risk factor for MI^35^, and (4) HT as a risk factor for atrial fibrillation (AF)^36^ (Supplementary Table 8, Supplementary Figure 4). We chose to assess direction of PGS-associated effect rather than overall heterogeneity of effect sizes due to the more dramatic inferential implications and because of systematically smaller sample sizes of secondary traits than primary traits (Supplementary Figure 5). By restricting PGSs to those directly developed for each primary condition (i.e. risk factor or exposure) that passed FDR<0.1 in *continuous* PGS transformation, we asked if it is more likely for GSGs to exhibit wrong association direction in the secondary outcome (i.e. flip-flops) when PGS heterogeneity is higher.

We found that the number of GSGs exhibiting flip-flops of PGS effect in secondary traits, as opposed to expected direction of risk increasing, is borderline significant with higher heterogeneity of PGS in primary trait, when pooled across all exemplary trait pairs while controlling for the four types of trait pair (beta=0.758, p=0.051, Figure 3c). The same regression test within each trait pair was not significant, except in T2D-MI (beta=1.195, p=0.031). A contrasting set of scores that explicitly highlights this observation (Figure 3b) is provided by a homogeneous T2D score (PGS000330^37^, I^2^ = 0) and a heterogeneous one (PGS003091^38^, I^2^=0.30). The homogeneous PGS showed a consistent risk relationship between T2D and MI for all GSGs, as expected. Conversely, the heterogeneous PGS showed a flipped, protective effect (i.e., increased risk of T2D with lower MI risk) in AFR and CSA (Figure 3c). This evidence is additionally mirrored by a related observation of heterogeneous PGS associated with more GSGs showing opposite association direction between primary and secondary traits (Supplementary Figure 6)

### Inference of features contributing to prediction accuracy and portability

To facilitate downstream PGS utility or guide future PGS development, we aimed to identify features that result in more accurate or portable PGS prediction. To do so, we performed separate multivariable regression analyses, or specifically, hierarchical linear mixed model (HLMM) for two outcomes: trait–PGS prediction accuracy (measured by incremental AUC) and ancestry heterogeneity (measured by across-GSG I^2^). Here HLMM was used to account for PGS and phecode as random effects while both have a nested structure: multiple PGSs can be developed under the same phenotype, and multiple phecodes can be under a parent phecode (e.g. “Alzeimer’s Disease” phecode 290.11 is nested within “Dementias/Unspecified” phecode 290.1). For each outcome, analyses were performed under both PGS transformations separately: *continuous* and *thresholded*. These HLMMs include 38 predictor variables spanning characteristics of the PGS and its constituent variants, the GWAS and training datasets, the test dataset, and phecode domains (Figure 4, Supplementary Figure 7, and Methods). Because weighted average of AUC can be driven by an overly large sample-sized GSG, we additionally computed a non-EUR-like incremental AUC across the rest of GSGs.

**Figure 4.**
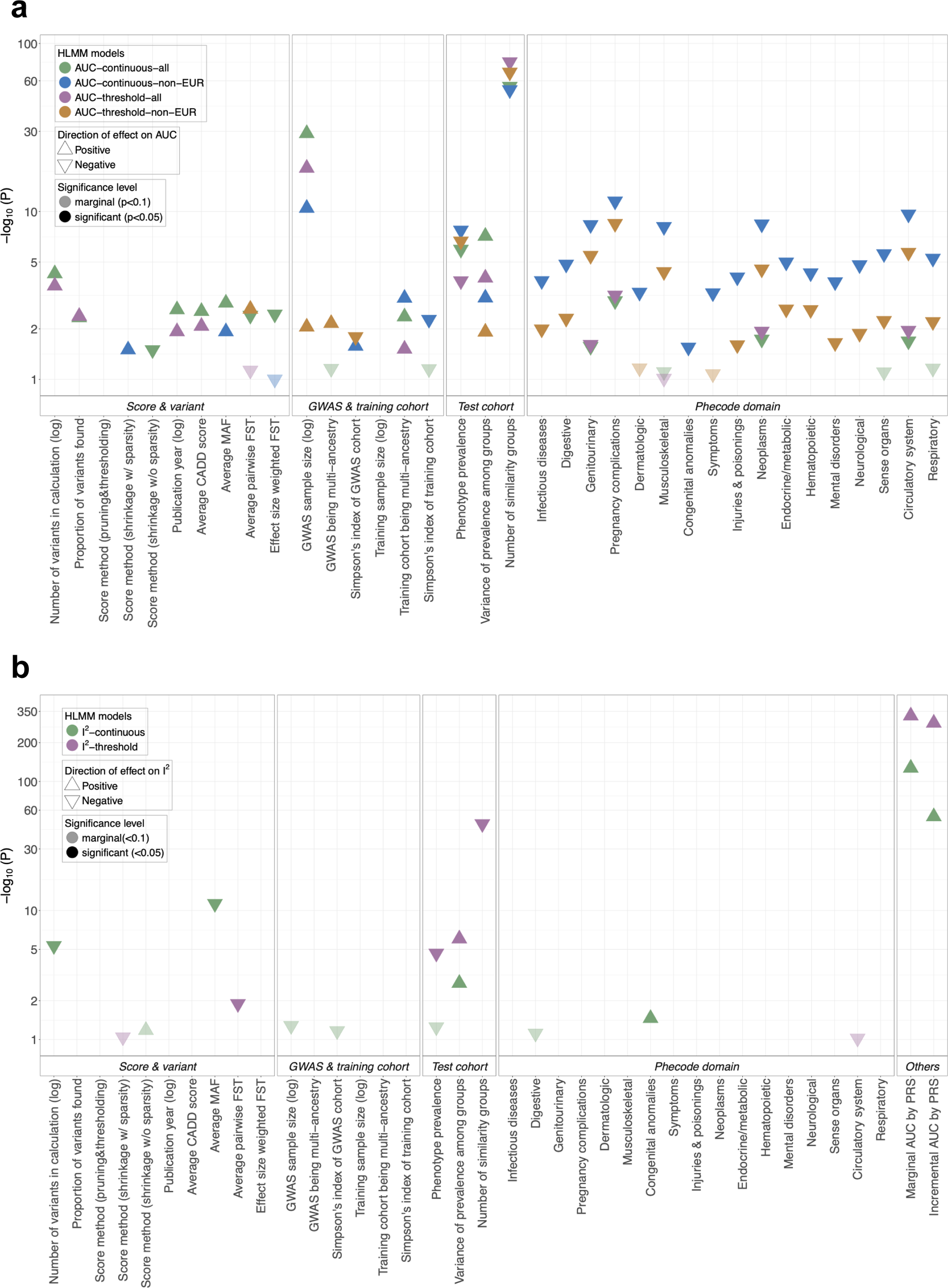
Multiple features associated with PGS prediction accuracy and cross-ancestry heterogeneity. **a**, Prediction accuracy, represented by incremental AUC, are associated with various features in scenarios of using continuous or threshold PGS measure, in all GSGs or those excluding EUR-like. **b**, Inverse portability, measured by heterogeneity metric of I^2^, are associated with various features in scenarios of using continuous or threshold PGS measure. Only significant (p<0.05) and marginally significant (p<0.1) features are shown in the plot, with up and down arrows depicting direction of each feature’s effect on incremental AUC or I^2^. Note that some covariate columns do not have any datapoint shown as they are not (marginally) significantly associated with the response variable.

Based on the above criteria, our framework consists of six feature-inferring HLMMs, including four explaining PGS accuracy: *AUC-continuous-all*, *AUC-threshold-all*, *AUC-continuous-non-EUR*, and *AUC-threshold-non-EUR*; and two explaining PGS portability: *I^2^-continuous*, *I^2^-threshold*. Below, we highlight the major trends to characterize pertinent features, and the full set of statistical associations per feature per HLMM are available in Supplementary Tables 9 and 14, with the significant associations shown in Figure 4.

### i. Cohort characteristics improving PGS prediction accuracy are subject to choice of PGS transformation and whether EUR-like is included

We observed a set of features consistently accompanying higher prediction accuracy across all four HLMMs for outcome of incremental AUC: larger GWAS sample size, lower phecode prevalence, higher variance of phecode prevalence among GSGs, and fewer number of GSGs in CCPM (Figure 4a, Supplementary Table 9).

However, this is not always the case for all associated features, as we identified discordant trends among HLMMs, particularly between settings whether EUR-like GSG was included or not. These contrasting features include types of PGS development methods, among-GSG genetic distance reflected by PGS-constituent variants (i.e. F_ST_-related features), and continuous measurement of sample diversity in original GWAS. Importantly for the last, we even observed contrasting effect on prediction accuracy between dichotomous (i.e. multi-ancestry or not) and continuous measure (i.e. Simpson’s index) of ancestry composition: in *AUC-threshold-non-EUR*, Simpson’s index in GWAS samples was associated with lower accuracy (beta =-8.66e-4, p=0.0161), while the multi-ancestry GWAS was associated with higher accuracy (beta = 7.55e-4, p=6.95e-3). The differences in feature importance for HLMMs illustrate that post-scoring interpretation, including how a PGS is transformed, and the test dataset ancestry composition, can influence patterns of important predictors.

We followed up with the contrasting effects between two measures of GWAS sample diversity and found that this could be partially interfered by a non-linear effect from Simpson’s index. By adding quadratic terms of Simpson’s index in GWAS and training datasets, we observe positive associations of those with incremental AUC (beta = 4.56e-3 and 0.760, p=0.0226 and 0.0380 for quadratic Simpson’s index in GWAS in *AUC-continuous-non-EUR* and *AUC-threshold-non-EUR,* respectively; beta =1.86, p=1.85e-3 for quadratic Simpson’s index in training in *AUC-threshold-non-EUR*; Supplementary Table 12).

Additionally, we found not all feature measures were intuitive to interpret, and conducted auxiliary analyses to give these findings deeper context. One such observation was lowered accuracy with ridge-like methods found in *AUC-continuous-all*. Prior work had indicated ridge-like methods show improved performance in certain settings ^15^. Given that score polygenicity itself was associated with higher accuracy in the same HLMM (beta=3.64e-5, p=5.43e-5), we speculated that the unexpected association direction with ridge-like methods type resulted from a tradeoff between the number of variants and prediction accuracy for ridge-like scoring methods. To test this, we replaced the four-level method category by an indicator of sparsity in method type (Supplementary Table 10) and included the interaction between number of variants*sparsity to test for the tradeoff effect in *AUC-continuous-all*. We did not identify a significant effect from either sparsity or the interaction (p=0.383 and 0.993, respectively, Supplementary Table 11). We also conducted the same test by restricting PGSs to those only using penalized methods (i.e. ridge-like or lasso-like), where number of variants is higher end than GWAS-hits or P&T. Similarly, these remained not significant (p=0.784 and 0.353, respectively, Supplementary Table 13).

### ii. Scattered features of PGS-constituent variants and test cohort are found to impact PGS portability, instead of score development

Different for the outcome of prediction accuracy, we did not observe “flip-flop” associations between HLMMs for the outcome of cross-GSG heterogeneity (as a qualitative reverse for portability) (Figure 4, Supplementary Table 14). Although in both portability HLMMs, heterogeneity was associated with accuracy, including both incremental and marginal AUC by PGS (beta = 3.06 and 1.72, p= 7.88e-128 and 3.36e-54 in *I^2^-continuous*, and beta = 1.02 and 5, p=6.94e-285 and <4.94e-324 in *I^2^-threshold*). It is likely that a higher AUC increased the sensitivity of the I^2^ by narrowing effect size confidence intervals (Supplementary Note). Most of other features with better portability reflect variant characteristics (e.g. numbers, MAF, F_ST_) or related to CCPM GSGs (Figure 4b, Supplementary Table 14). While most were expected, we noticed an association between higher number of GSGs in CCPM and lower heterogeneity (beta =-0.0429, p=1.09e-47) in the *I^2^-threshold* HLMM. We wanted to ensure our observations were not influenced by including all groups, even those with more modest sample sizes. We re-tested our finding by only calculating I^2^ within the three largest GSGs with N>2,000 (EUR-like, AMR-like, AFR-like). This subgroup analysis changed the direction of effect and but were only suggestively significant in *I^2^-continuous* (beta = 5.29e-3, p=9.42e-2). The trends are still consistent in the *I^2^-threshold* HLMM for the 3-population sub-analysis, albeit somewhat attenuated (beta=-3.41e-2, p=3.25e-29) (Supplementary Table 15).

## Discussion

In our study we provide a comprehensive analysis of existing polygenic risk scores and associations across the entire phenome. We systematically evaluated prediction performance for each score and its association with each trait in terms of both accuracy and portability (homogeneity of accuracy) across ancestries from multiple GSGs in a large biobank. We show a large number of associations across most phenotype domains, particularly for codes for common diseases and traits. Heterogeneity in performance across genetic similarity groups is highly variable across scores and traits and is affected by choice of PGS unit. This can have implications for any downstream analysis, particularly clinical and translational investigations that may be more interested in thresholding or identifying a group at higher genetic risk of a condition.

Specifically, we highlight the consequences of adapting PGS with greater heterogeneity into secondary trait analyses like MR, to demonstrate that low portability can propagate into larger errors with the second trait. Importantly, we characterized both accuracy and portability of PGS performance using a broad array of features representing qualities of the score, training and test data, and features of our test setting in CCPM, which was all out-of-sample. Our study offers a description of features associated with improved performance, as well as trends through time as GWAS have grown in size and have understood the importance of capturing multi-ancestry designs and settings.

Our phenome-wide association results attest to the great potential of PGS in phenotypic prediction for clinical and translational applications, echoing observations from previous trait-specific PGS investigations in other studies ^2,3^. The variability in statistical significance among traits and domains likely has multiple attributes: first, common diseases with large prevalence (e.g. metabolism related outcomes) result in higher statistical power in both the original GWAS and the test dataset. Second, phenotype domains do not contain a uniform number of scores. The ones with greater research attention, such as cancer, have more scores developed (Figure 2a). Third, the varying genetic architecture across traits can itself impact both statistical power in GWAS and subsequent PGS applications. As a retrospective study, we note the most significant scores are a composite of high-performance scores and high-prevalence traits, however, we release all summary statistics to enable researchers to prioritize scores based on our analyses.

Despite the large number of significant scores, the challenge of portability across GSGs continues to manifest even in scores developed recently. We see this evidence in the extent of heterogeneity between groups quantified in our study, whether driven by genetic differences driving trait differences or, more likely, residual population stratification. Instead of capturing trait-specific genetic profiles through high-heritability pathological pathways, PCA of this large collection of PGSs indicated that the leading drivers of score dimensions align with both global and intra-European variation. This reflects both population structure in PGS ^11,39,40^, and the ancestry-biased training common of most scores.

To move away from discrete population labels where possible to reflect the continuous nature of human genetic diversity ^41^, there has been recent effort in the field proposing *post hoc* calibration of ancestry in calculated scores to address population stratification ^4,8,42–44^. These methods standardize a score’s distribution across ancestry by removing the estimated contribution from individuals’ ancestry on both the mean and variance of score values. However, we decided not to take this approach in this study, due to (1) the study’s intention to evaluate PGS under the most common ancestry-stratified practices via GSG-specific association testing, (2) the retrospective study design with ancestry adjustment provides a similar adjustment via multiple regression per trait, and (3) the concern of a one-size-fits-all adjustment approach that could be difficult across a phenome-wide examination ^27^. As an example, skin pigmentation/tanning and related outcomes, such as melanoma, can be exceedingly population-stratified with potential causal variants highly segregating among ancestries^45^, where a standard calibration method could attenuate any true genetic predictive values. When it comes to specific traits of interest, however, the use of ancestry calibration still has great promise if the genetic architecture of the trait is largely shared across ancestry in test populations, as is common for highly polygenic traits. Additionally, evaluation of the fit of scores before and after calibration can likely provide more information on how best to incorporate these approaches.

There has been growing interest in understanding PGS performance in large datasets around the world. Recently, Kolosov et al. provided platform-based resources that focused on accuracy evaluation of over 2,500 PGS models in the relatively ancestry-homogeneous FinnGen cohort, offering important empirical estimates of prediction performance of a bulk of existing PGSs ^17^. In this study, we released a compendium of score attributes and phenome-wide summary statistics focused on the question across six genetic-similarity-groups in the biobank at CCPM. We placed additional emphasis on evaluating cross-ancestry portability alongside accuracy without assuming goodness-of-fit current ancestry calibration approaches. By further exploiting PGS-trait associations, we enabled inference of both portability and accuracy by identifying correlates of score methodologies, genomic features, and cohort characteristics based on a contemporary snapshot of available scores. Such trends can be hard to capture using conventional benchmarking designs alone.

We note that while we attempted to identify predictors across a range of features reflecting both the genome and study design, the features that emerge above statistical significance are merely a limited set of potential sources of score information. The models, while developed to reflect the correlation structure present in traits and scores, could reflect additional confounding or indirect associations that are difficult, if not impossible, to fully resolve at this stage. Many of the performance-enhancing features identified through HLMM are consistent with expectations, such as larger size of scores, newer genome-wide models, and larger GWAS study size.

Leveraging multi-ancestry GWAS or training datasets was associated with greater accuracy in general and marginally with attenuated heterogeneity when measuring *continuous* PGS units, replicating the conclusions from our prior study ^46^ and observations from a selection of quantitative traits ^13^. However, we caution that in this dataset increasing the number of ancestries in the GWAS or training panel does not necessarily improve all measures of performance in all situations: while this trend was confirmed by accuracy improvement in multi-ancestry settings than single ancestry, the opposite was observed in quantitative measure of diversity in some of the accuracy-evaluation HLMMs. When designing a PGS with a specific purpose or target population in mind, it would be worth examining whether closer matching of ancestry composition at finer granularity can help address this issue.

Additionally, we did not find a straightforward interpretation for the association of higher pairwise F_ST_ with attenuated heterogeneity and increased accuracy in some of HLMMs (but not all), as one may instead expect the opposite, if higher F_ST_ were to lead to greater heterogeneity. We speculate that the F_ST_ measure calculated from PGS variants here may reflect multiple underlying attributes, including but not limited to the proportion of variants that are ancestry informative in a score model, whether traits are structured among tested GSGs, and whether stratifying PGS with a fixed threshold for all groups (i.e. top decile in this study) weakens performance in score models that do not capture ancestry informative alleles. Addressing this question will likely benefit from more detailed investigations into the mechanism of score development and variant selection under more controlled settings, which is beyond the scope of the present study.

Our models also found surprising associations that may be due to inevitable limitations of metrics. For example, I^2^ can be deflated among groups with larger statistical uncertainty or smaller sample sizes. The observed discrepancies of I^2^ between different PGS units (e.g. per-SD or top decile over the rest) could result from the inherent uncertainty present in individual-level scores particularly when thresholded ^47^. Leveraging per-individual measures of uncertainty could help address this ^47^ but are not simple to store in static repositories and could be difficult to interpret for downstream purposes, particularly in clinical and translational settings.

Overall, our study shows both the potential and the challenges of applying PGS for various health outcomes through evaluations across a high dimension of scores, phenotypes, in a large-scale biobank with participants from diverse backgrounds. We provide a repository of 5,747,366 score-trait associations, including 371,196 continuous and 206,840 thresholded score-trait pairs that meet our false discovery rate criteria for statistical significance, enabling researchers to apply this in downstream analyses and provide a complement to existing public resources such as UK Biobank^48^ and the All of Us Research Program^49^. There are still considerations of ancestry-aware approaches and caution is needed in score choice and interpretations. However, as part of this work we demonstrate that through this large sifting exercise we can identify numerous scores that enrich for ancestry portability, making them higher priority for downstream applications. Overall, we hope this resource provides relevant information to evaluate accuracy and portability, along with the efforts from country-based and international consortia, to provide greater clinical and translational opportunities for polygenic scoring.

## Methods

### Regulatory Approval

The biobank at the Colorado Center for Personalized Medicine study data and protocol, including primary consents, was approved under Colorado Multiple Institutional Review Board #15–0461 and 20–2289, with additional approval for research operations via the CCPM Executive Committee.

### CCPM Biobank freeze3 dataset

The biobank at the Colorado Center for Personalized Medicine (CCPM Biobank) was jointly developed by the University of Colorado Anschutz Medical Campus and UCHealth, with the majority of participants from the Rocky Mountain Region. The CCPM Biobank integrates research and clinical resources, comprising electronic health records (EHR), genotype data, and more ^25^. Freeze 3 version of the CCPM Biobank consists of 73,346 participants whose genetic data were generated by a custom Infinium Multi-Ethnic Global Array SNP chip (MEGA, Illumina, Inc.), or whole exome sequenced in partnership with Regeneron Genetic Center, and subsequently imputed through TOPMed imputation server.

Details regarding sample collection, data generation and imputation, quality control, relatedness inference, PCA and genetic ancestry assignment for our dataset are described in *CCPM Freeze3 Materials and Methods* ^26^ and the description of the biobank ^25^.

We used PheWAS package^29^ in R to map ICD-9 and ICD-10 codes from CCPM participants to phecode based on phecode definition v1.1, where cases were considered if minimum count was 2. Phecode domain information was later assigned using the phecode definition v1.2 at https://phewascatalog.org/phecodes.

### Retrieving and applying score models from PGS catalog to the biobank

We obtained 4,742 score weight files (developed for 680 mapped EFO traits) from the PGS catalog (https://www.pgscatalog.org)^24^, of which 13 have CCPM Biobank as one of their training cohorts and thus were excluded from this study. We then used our PGS harmonization and calculation pipeline, ESCALATOR (https://github.com/menglin44/escalator, Supplementary Figure 1), to apply the weight files to the imputed dataset of 73,346 individuals for score calculation. Among them, 147 score models did not specify genome build information or build information is not recoverable, 579 did not have enough effective columns for our pipeline to generate correct scores (e.g. missing non-risk allele codes), 5 had no variants overlapping with the biobank imputed dataset, 4 had 0 as weights for all variants and were effectively non-informative. This resulted in 3,994 PGSs available for subsequent analyses. Note: while the terms PGS, PGS and GRS are all common, for consistency we use the term PGS across all analyses.

### Ontology-based PGS category

We obtained the PGS category information from the trait category metadata of PGS catalog (https://www.pgscatalog.org/browse/traits/), which provides assignment of an ontology identifier (EFO ID) to one of the 16 exiting PGS categories. We then combined this information with mapped trait ontology of each PGS from https://ftp.ebi.ac.uk/pub/databases/spot/pgs/metadata/pgs_all_metadata_scores.csv. There were eight EFO labels with missing or incomplete assigned categories at the time of our analysis, and we manually assigned them based on their close matching traits and assignments, including: fat body mass and hip circumference classified into “Body measurement”; waist circumference, serum albumin measurement, leukocyte count classified into “Other measurement”; unipolar depression and Disorder of bilirubin metabolism and excretion classified into “Other trait”; alcohol consumption measurement classified into “Biological process”. There were additional 46 ontology identifiers where 628 PGSs were developed for (or 338 PGSs with no additional assigned ontology) having no assigned PGS categories, further grouped as “unassigned”.

### Dimensional reduction on PGSs

We performed principal component analysis (PCA) on all 3,994 PGSs. Analogous to the concept of pruning out related SNPs in LD, we pruned out highly related PGSs that are designed for the same phenotype based on EFO labels. For each EFO phenotype with multiple PGSs, we selected the PGS that had the lowest p-values available across phenome associations, and we excluded PGSs designed for multiple phenotypes to avoid the ambiguity in the phenotype assignment (excluded n=475). This yielded 518 “independent” scores across all biobank individuals. We then performed PCA with 518 variables using *prcomp* function in R on the pruned score by unrelated individuals.

### Phenome-wide associations across all PGSs

We conducted the atlas of all-by-all association tests between each phecode and each PGS within each of the six similarity groups. We used the logistic firth algorithm now implemented in PLINK 2.0 (www.cog-genomics.org/plink/2.0/)^50^ to facilitate unbiased estimate in phenotypes with imbalanced case and control counts. The model corrected for age, sex, genotyping batch, and top ten principal component analysis (PCA) vectors. To make continuous PGS values to be accepted by PLINK for association testing, we rescaled the PGS values to a dosage range between 0 and 2 that aligned with diploid genotyping dosages, and scaled back the reported summary statistics (e.g. effect size) to that reflecting 1 SD increase of score values. When taking PGS as its thresholded form, we converted the PGS values to binary form thresholded at top decile within each similarity group. The associations were only run in phenotypes that have more than 10 cases per test. When reporting results, highly significant associations that were beyond the limit of the smallest positive subnormal number in double-precision floating-point format from IEEE standard (i.e. p<4.94e-324) were truncated to aid in interpretability.

### Meta-analysis and significance threshold

We meta-analyzed the phecode-PGS associations across similarity groups by using METASOFT (http://genetics.cs.ucla.edu/meta/index.html), separately for taking PGS as its continuous unit and its thresholded unit. We also separately meta-analyzed when excluding EUR-like group for part of the subsequent HLMM analysis. As METASOFT accepts standard errors other than p values, yet the standard errors from association tests were not under a standard Wald test model, we converted effect sizes and p values from Firth to a pseudo standard error under Wald test model assumption for METASOFT input. As this study focuses on evaluations of PGS across ancestries, we only kept associations that have been run in at least more than two similarity groups, ending with 5,747,366 phecode∼PGS association pairs (and 4,625,052 when excluding EUR-like).

As METASOFT provides a more powerful estimate of random model where the effect size under the null assumption is not zero (Han and Eskin, 2011), we extracted p values from both fixed and the new random model and report the association p values based on estimated heterogeneity:

When I^2^ is relatively homogeneous (<0.2), we take the fixed model p values to report; When I^2^>0.2, the Han and Eskin’s random model p values. Due to the high relatedness among phecodes and PGSs developed under the same trait, we adopted FDR<0.1 as our significance threshold, ending with 371,195 and 206,840 significant associations when taking PGS in continuous and thresholded unit, respectively.

In addition to p values and effect sizes, we additionally extracted information of similarity groups and the number of them, I^2^, and *τ*^2^ (DerSimonian-Laird estimator) per phecode∼PGS association pair from meta-analysis.

### Counting flipped associations in known causal trait pairs

We counted the number of GSGs with unexpected association direction, or flip-flops, between PGS designed for a primary exposure and its corresponding secondary outcome phenotype. By expectation, PGS exhibiting a negative, or protective effect with the secondary trait is defined as a flip-flop. However, we did notice a few models having effect of PGS flipped in design that it exhibited a strong protective effect even in the primary exposure. Thus, for PGSs that showed a protective effect in over 5 GSGs in the primary exposure, we counted the number of GSGs with positive association with the secondary trait as its number of flip-flops. Separately, we counted the number of GSGs having PGS-association flip between the primary and the secondary conditions.

### Area under the curve (AUC) of associations

AUC of each phecode∼PGS pair was calculated using *pROC* package in R, in three different generalized linear models (GLM): (1) a full model including all covariates (age, sex, batch, top 10 PCs) and PGS. (2) a model including non-PGS covariates, and (3) a model including only PGS. Incremental AUC was calculated as the subtraction of AUC values between (1) and (2). Calculations were performed in unrelated individuals in each similarity group. Then overall weighted average AUCs across groups were calculated over per-group AUC weighted by their sample sizes.

### Features related to training datasets

Some features are directly extracted from the summary meta datasheet from PGS catalog, including the year of publication, and original phenotypes scores were developed for as in standard EFO categories. Some other features were further refined based on the primary information provided in the dataset –

(1) Sample size of GWAS and training cohorts

Some scores are reported with multiple entries of GWAS sample sizes (with “Source of Variant Associations (GWAS)” under the stage information), often in situations like meta-analysis or combining multiple GWAS summary statistics for score development. If such a case is present, the sample size of GWAS is taken as the sum of those entries;

Otherwise, the single entry of GWAS sample size. Similarly, sample sizes reported with “Score Development/Training” under the stage information are considered training sample sizes, and are combined if multiple entries appear for the same score.

(2) Ancestry diversity within GWAS and training dataset

By extracting the reported ancestry distribution for GWAS and training stage from the “score” table of the meta sheet, we first assigned a binary description of GWAS or training datasets being multiple ancestry. If the entry described one single ancestry as 100% of source, this was translated to multiple ancestry; if not (e.g. multiple ancestry with partial percentages that summed up to 100%), this was translated to single ancestry; blank entries were considered not available (“NA”).

Then, for scores with available ancestry information, we further calculated Simpson’s index to better capture the extent of ancestry diversity as follows:

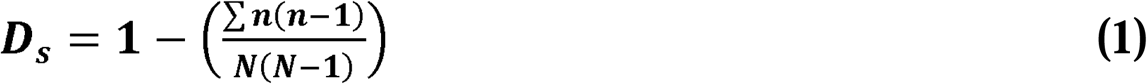

With D_s_ being the Simpson’s index, n is the number of individuals within a reported ancestry genre back calculated from the total sample size and the percentage, is the total sample size.

(3) Type of PGS development methods

While there has been an extensive description of development methods or software used in each score, we categorized them into four main groups based on the nature and similarity of these methods: (1) GWAS significant hits, by stringently picking the signals above the genome-wide significance threshold, and the signals are independent in terms of linkage disequilibrium (LD); (2) Pruning and thresholding (P&T), by including independent hits that are above a customized, often relaxed p-value threshold after LD pruning or clumping; (3) PGS score with extensive sparsity, often by methods that estimates a posterior effect size distribution in consideration of LD (e.g. “beta-shrinkage”) that essentially excluded variants that are unlikely to contribute non-zero effect sizes to the model, such as lassosum; (4) PGS score with beta-shrinkage methods, but no sparsity steps were applied, such as popular LDpred2, where this type of methods often leave with score models of large number of variants (typically around or above 1M).

We note that there are a few scores adopting some late methods in the fashion of combining existing score models, instead of starting from GWAS summary statistics, such as PGSmix or ensemblePGS. Essentially, there is a large heterogeneity of method nature within such type due to the uncertain variation of input score models and we find is not wise to individually back track nor possible to characterize. We excluded scores from being assigned with a method type.

(4) Number of variants in the score model

To reflect both the score size and the overlap of variants between the model and the test dataset, we included both features of the number of variants of a score found and used in the CCPM Biobank, and the proportion of such as compared to the original number of variants in the model.

### Features related to the test dataset

(1) Phecode prevalence in CCPM biobank

This includes the average prevalence of the phecode, and the variance of such prevalence among the similarity groups.

(2) Phecode domain

The definition and the grouping information of phecode (or disease domain), is obtained from the PheWAS catalog (https://phewascatalog.org/phecodes)

(3) CADD scores

We incorporated a broadly applicable annotation of the genetic variants used to derive each PGS by analyzing the Combined Annotation Dependent Depletion (CADD) scores across all 3,944 models extracted from PGS Catalog. A genetic variant’s CADD score provided an objective criterion to estimate the degree to which single nucleotide variants are functional, deleterious, and causal. We downloaded the annotation file from https://cadd.gs.washington.edu/download and then subset this annotation to the set of variants present in the imputed CCPM Biobank genotype data. The CADD scores for these 18.3 million variants were then merged with each of the 3,944 PGS weights files in order to extract the distribution of scores per training dataset. For downstream analyses, the mean CADD score was used to quantify the average functional relevance of variants in each PGS.

(4) Number of similarity groups

As not all similarity groups have >10 cases for a condition to be included in the meta-analysis, we summarized the numbers of similarity groups that were effective part of each meta-analysis association.

### Features related to both training and test dataset

(1) Score divergence among groups as proxied by F_ST_

For each phecode∼PGS pair appeared across similarity groups, we calculated the genetic distance reflected by the score variants as both (1) the mean F_ST_ over all variants used in the score calculation, and (2) a weighted F_ST_ to weight in the differential contribution from risk effects across variants.

The first was done in the standard ratio of average as in Hudson’s F_ST_ estimator (Hudson et al., 1992; Bhatia et al., 2013), between pairwise similarity groups that contributed to meta-analysis, denoted as here. The second was calculated as the weighted sum over per-variant *F_ST_* by a within-score standardized risk effect, between pairwise similarity groups, denoted as.

More explicitly, we have the average *F_ST_* (Hudson’s estimator) and per-effect weighted *wF_ST_* respectively as

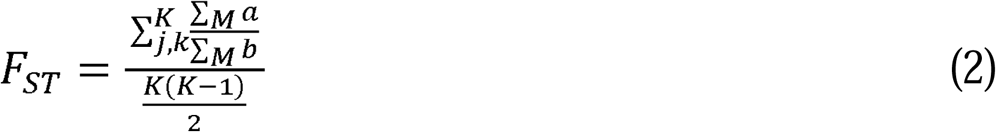

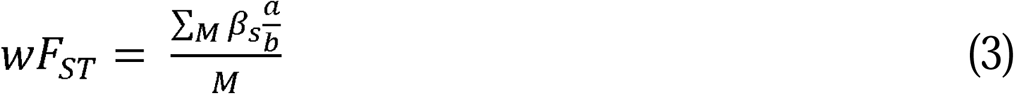

The intermediate terms and are defined as

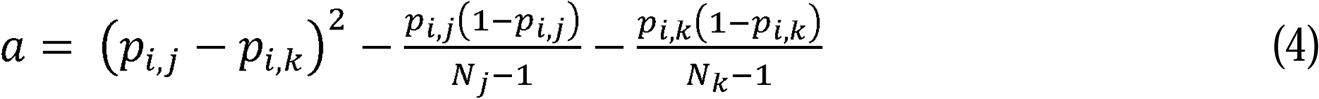

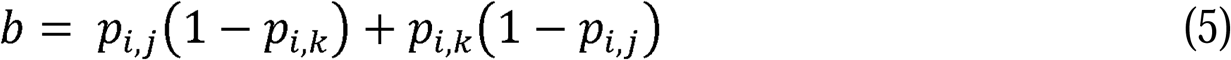

For all distinct integers 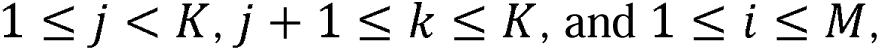 where K is the number of ancestry groups from which summary statistics contributed to meta-analysis in the association pair, and M the number of variants used for calculating the PGS.

Thus, the two F_ST_ estimates are defined as

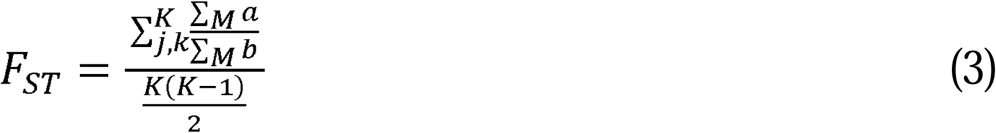

and

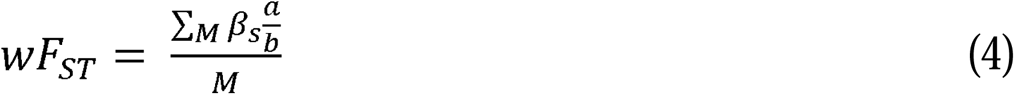

where is the standardized weight per risk variant.

(2) Average minor allele frequency (MAF) of a score

Commonality of alleles is associated with a broad range of information, including whether the variant carries estimated risk effect is prevalent in a population, and whether this variant tends more likely to be population-specific, given human population’s recent migrations with several major bottlenecks and subsequent latest expansion. To proxy the quality of interest, we recorded the average of MAF over variants used in each score.

### Hierarchical linear mixed model

Modeling I^2^ and incremental AUC (or beta effect of PGS in a full model, when PGS is taken as in a thresholded form) as response variables, we performed associations between response variables and curated feature covariates reflecting characteristics from the test and training dataset for each phecode∼PGS pair, via *lme4* (Bates et al., 2014) and *lmerTest* (Kuznetsova et al., 2017) packages in R. We model the random effects of repeated encounters of PGSs and phecodes, and hierarchical structures that account for the nested nature of phecodes (e.g. 250.11 of “type 1 diabetes accompanied by ketoacidosis” is nested under 250.1 of “type 1 diabetes”), and multiple PGSs developed for the same traits defined by EFO ontology identifiers. There were 475 PGSs that are assigned with multiple EFOs from their original phenotype(s): each PGS and its associated entries was duplicated per each EFO within the dataset in preparation of HLMM. For phecode∼PGS pair where the score is developed for multiple EFO phenotypes, we duplicated the entry assigned to a single EFO at a time. When modeling I^2^, we only included associations that passed FDR<0.1 significance threshold, as I^2^ are likely to be deflated among non-significant associations with large standard errors. When modeling incremental AUC or beta effect of PGS, we drew a matched set of associations from non-significant pairs to be combined with the significant set, to ensure a balanced representation of spectrum of prediction accuracy. Some models with the data being subset to certain criteria (in the sub-analyses) did not converge under the default BOBYQA optimizer in *lmer*, and were switched to Nelder-Mead optimizer.

## Data Availability Statement

We provide the full summary statistics of all scores by all phecodes, including those used for HLMM analyses at https://doi.org/10.5281/zenodo.22225045. The PGS harmonization and calculation pipeline, ESCALATOR, is available at https://github.com/menglin44/escalator. Visualization of top PGS-trait associations in CCPM: Web Overview of Multi-ancestry Polygenic scores in Biobank Traits (WOMPBAT) is available at https://wompbat.simonwalker.tech/.

## Acknowledgements

We appreciate the availability of the genetic data generated by the partnership between CCPM and Regeneron Genetics Center. EHR-linked data supported by the Health Data Compass Data Warehouse Project, healthdatacompass.org. We thank all the participants in CCPM biobank without whom this work would not be possible. We are grateful of the PGS resource made publicly available by the developers of PGS catalog. We thank the members of the CCPM PGSUIT working group, along with Eimear Kenny, Noah Zaitlen, and Valerie Arboleda for helpful conversations. This work was partially supported by Colorado Center for Personalized Medicine, and NIH grants U01HG011715, R01HG011345, and R01HL151152. The content is solely the responsibility of the authors and does not necessarily represent the official views of the National Institutes of Health.

## Author Contributions

ML and CRG conceptualized and designed the project with input from AEH and SR. NR, KRC, NP, JAS, MF, SC, HS, and CCPM provided the data. ML led the analyses with CHA and MF. ML and CRG interpreted data and results with feedback from AEH, SR, CHA, MP and LAL. SNKW and RML designed web visualization with input from ML and CRG. ML and CRG wrote the initial draft with input from CHA, SNKW, NR, KRC, MF, SC, HS, NP, LAL, MP, JAS, RML, AEH, SR.

