## Supplementary Figure for "The landscape of polygenic score prediction accuracy and transportability across the phenome in ancestrally diverse populations"

### Supplementary note: Choice of heterogeneity metric

Commonly used metrics to measure heterogeneity in genetic associations include Cochran's Q, $\tau^2$, $I^2$, and $H^2$. Among them, Q and $H^2$, while related to $I^2$, can be underpowered in small group numbers (in the case of this study, the number of similarity groups. We therefore compared $I^2$ and $\tau^2$ as potential candidates to quantify heterogeneity.

$I^2$ has a unique advantage in interpretability, as its range is restricted between 0-1. However, $I^2$ close to zero can be over-interpreted as support for homogeneity when estimates from groups have large statistical uncertainty. $\tau^2$ reflects a complementary measure of between-study variance. We compared the estimates of $I^2$ and $\tau^2$ of associations that passed FDR<0.1 to measure concordance between the two, particularly towards the low end, which would imply $I^2$ as a biased measure at lower effect sizes or greater standard errors. However, both metrics are output by METASOFT (we chose the DerSimonian-Laird estimator for $\tau^2$ , as the REML estimator failed to converge in many association pairs).

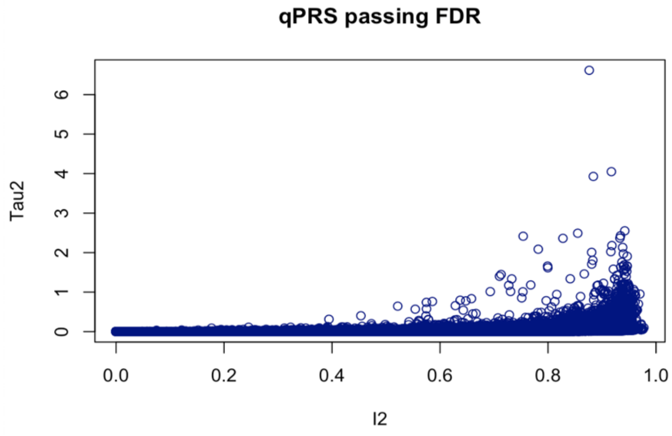


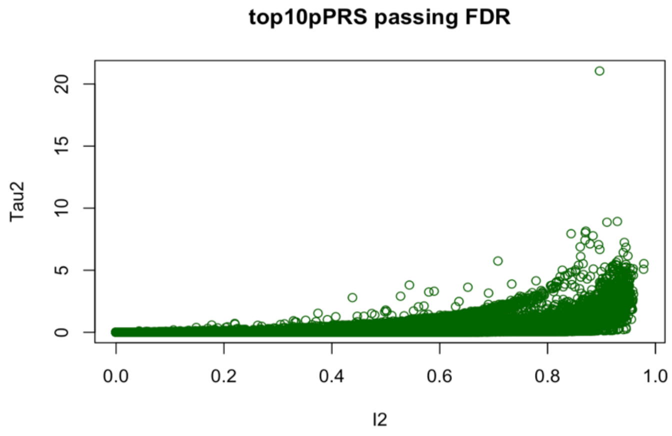

**Supplementary Note Figure 1. Comparison of $I^2$ and $\tau^2$ within the significant set of phecode-PGS associations**, when using continuous PGS (upper panel), and thresholded PGS (lower panel).

There is high correlation between the two measurements (Spearman's correlation coefficient = 0.978 and 0.984 for *continuous* and *thresholded* PGS, respectively), and we did not observe higher $\tau^2$ than $I^2$ at the lower end (Supplementary Note Figure 1). Based on this comparison we chose to focus on the more intuitive metric of $I^2$.

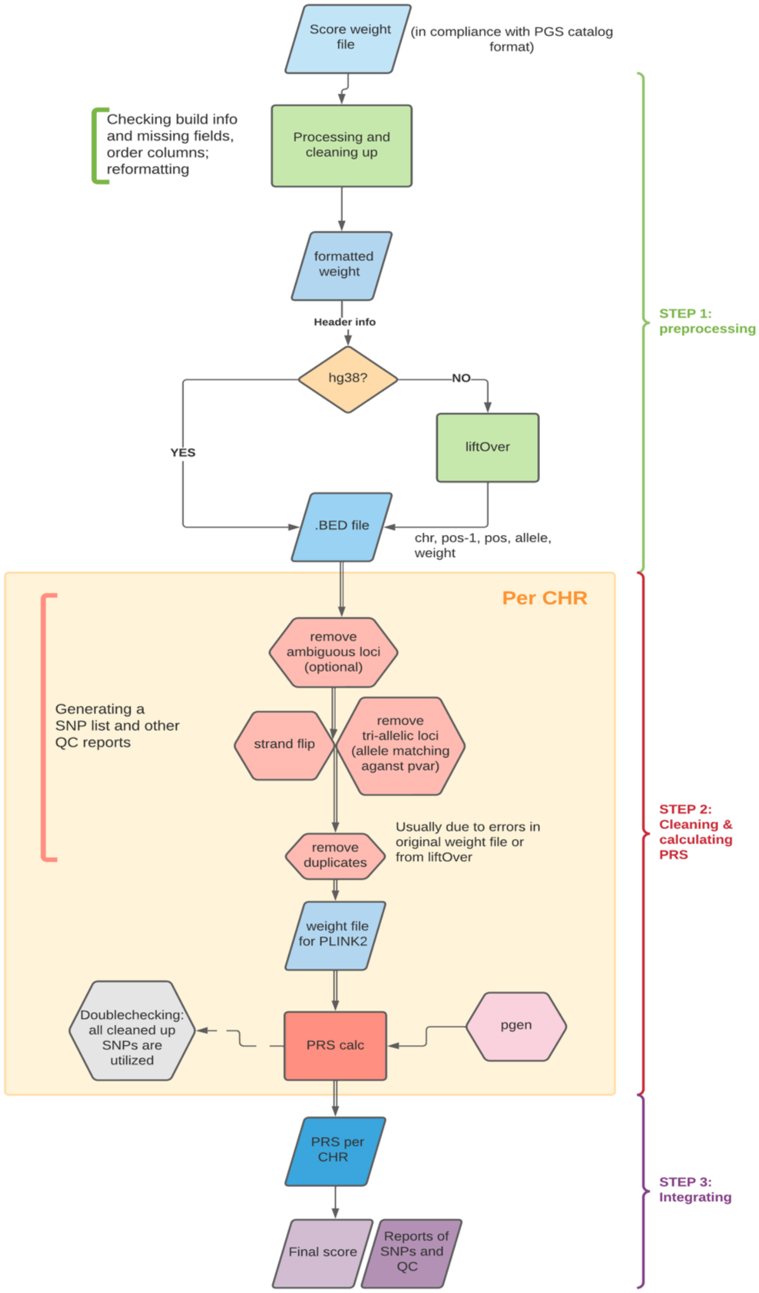


**Supplementary Figure 1. Scheme of ESCALATOR used to harmonize and apply PGS weights to CCPM biobank data.** Pipeline available at https://github.com/menglin44/escalator.

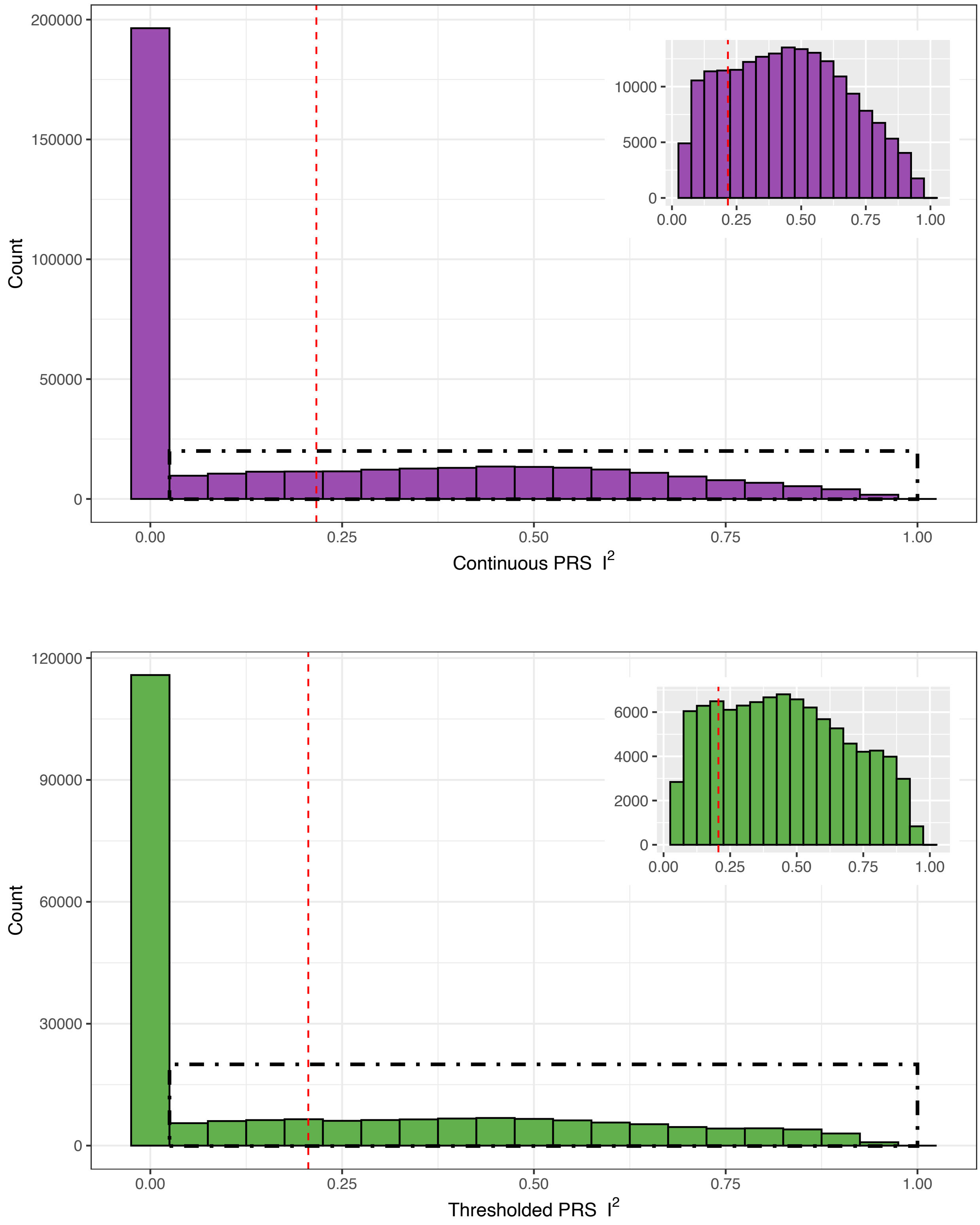


**Supplementary Figure 2. Histogram of $I^2$ in the significant set of associations** using *continuous* (upper panel) or *thresholded* (lower panel) PGS unit. Red dashed vertical lines denote the mean $I^2$, where the insets are the zoom-in of bins in the dashed rectangles.

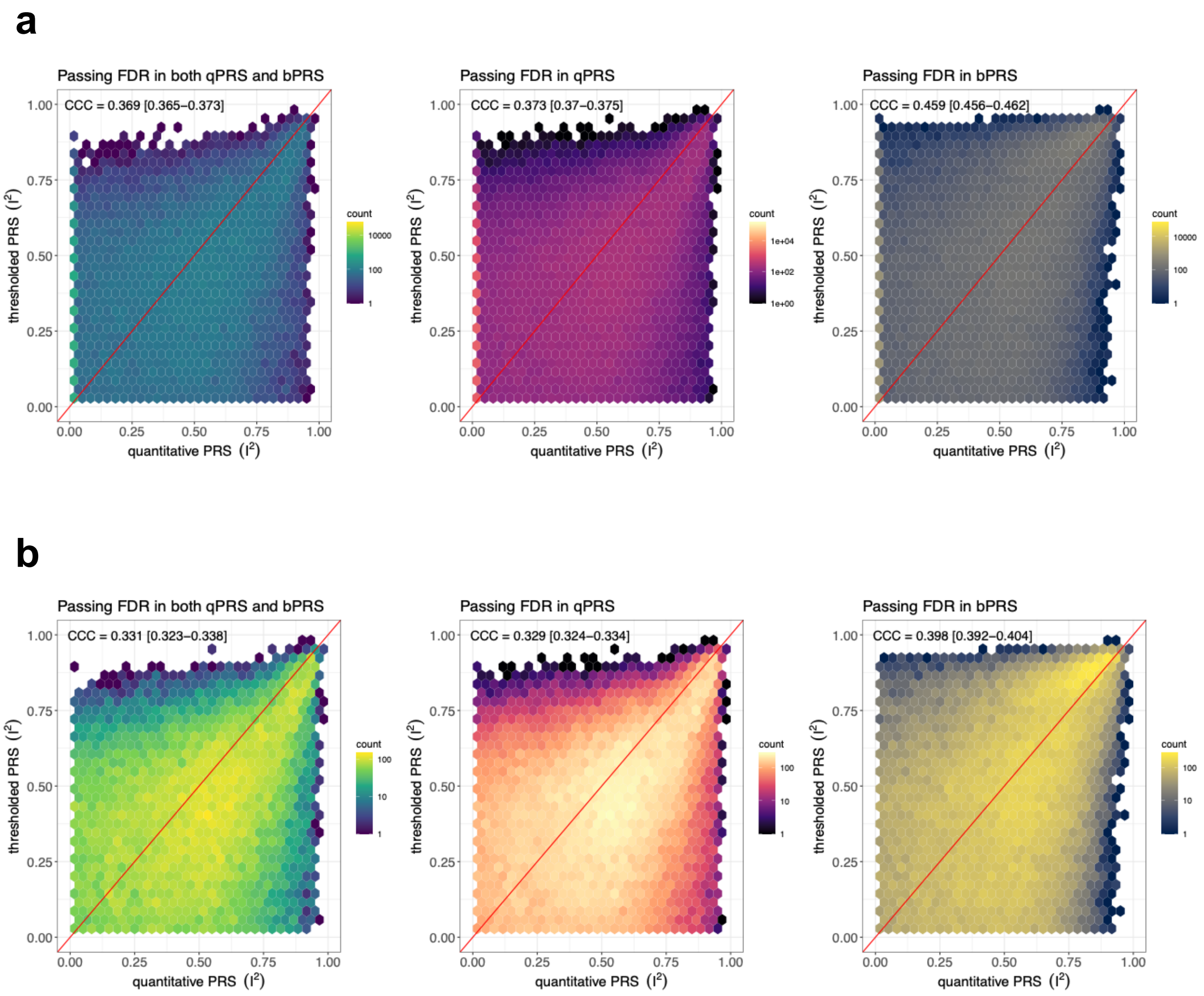


**Supplementary Figure 3. Correlation of phecode-PGS association $I^2$ between different PGS units**. **a**, $I^2$ measured using continuous PGS (x-axis) against that measured using thresholded PGS (y-axis) are plotted in hex bins for association pairs that pass the significance thresholds of both using continuous and thresholded PGS (left), and those passing that using significance threshold of continuous PGS (middle), and then of thresholded PGS (right). **b**, to examine if extreme low $I^2$ potentially affected by low statistical power in associations distort the correlations, the same correlations were mirrored by removing $I^2$=0 data points. Estimates of Lin's CCC [95% C.I.] are in the top of each panel.

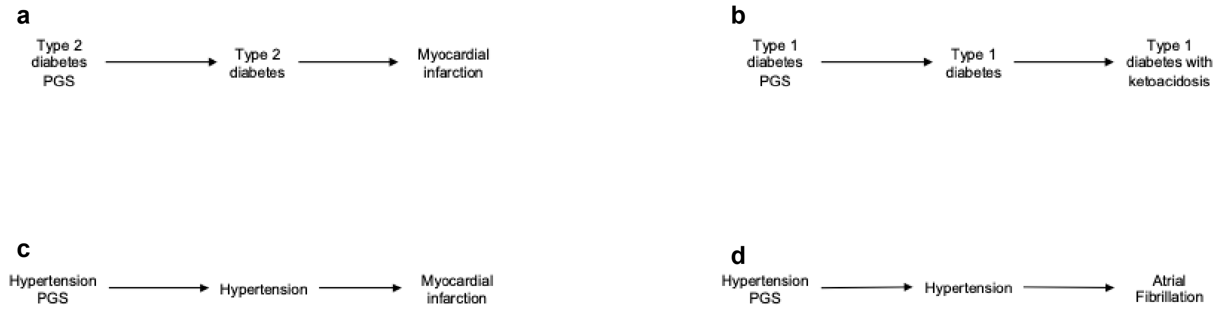


**Supplementary Figure 4. DAGs of four trait pairs with established causal relationships** tested in confirmatory MR tests for qualification of instrumental variables (IVs), under the hypothesis that an IV using heterogeneous performing PGS can propagate problematic inference under this kind of scheme.

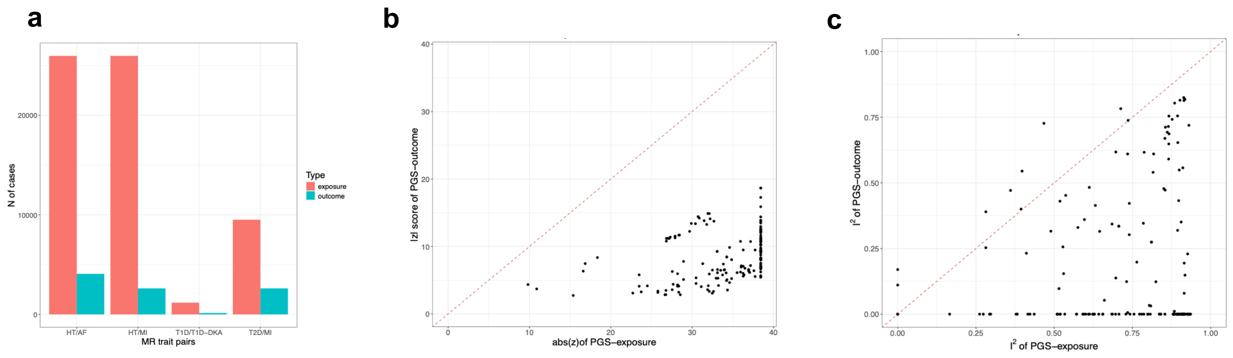


**Supplementary Figure 5. Case numbers and the impact on summary statistics in the selected primary-secondary MR trait pairs. a,** Across the four selected MR trait pairs, the number of cases in the primary trait (exposure) far exceeds that in the corresponding secondary trait (outcome). **b,** The absolute standardized effect size from PGS-trait association. Larger for exposure than corresponding outcome. Note: low p-values output as 0 from association summary statistics were converted to compatible double-precision float 4.94e-323 for computing z-score in this plot. **c**, $I^2$ in PGS-exposure is systematically larger than in corresponding PGS-outcome.

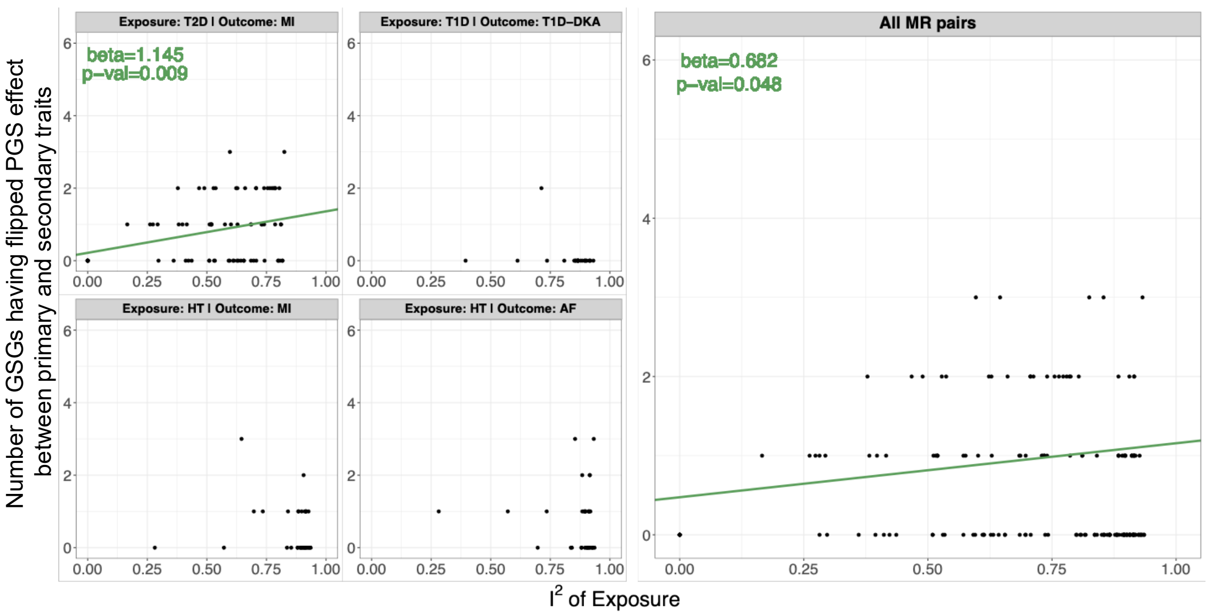


**Supplementary Figure 6. Number of GSGs having opposite PGS association directions between primary and secondary traits, against $I^2$ of PGS on the exposure trait**, within four selected MR pairs (left sub panels), and all pooled pairs (right large panel). Regression line and coefficients are shown in green if the regression is significant.

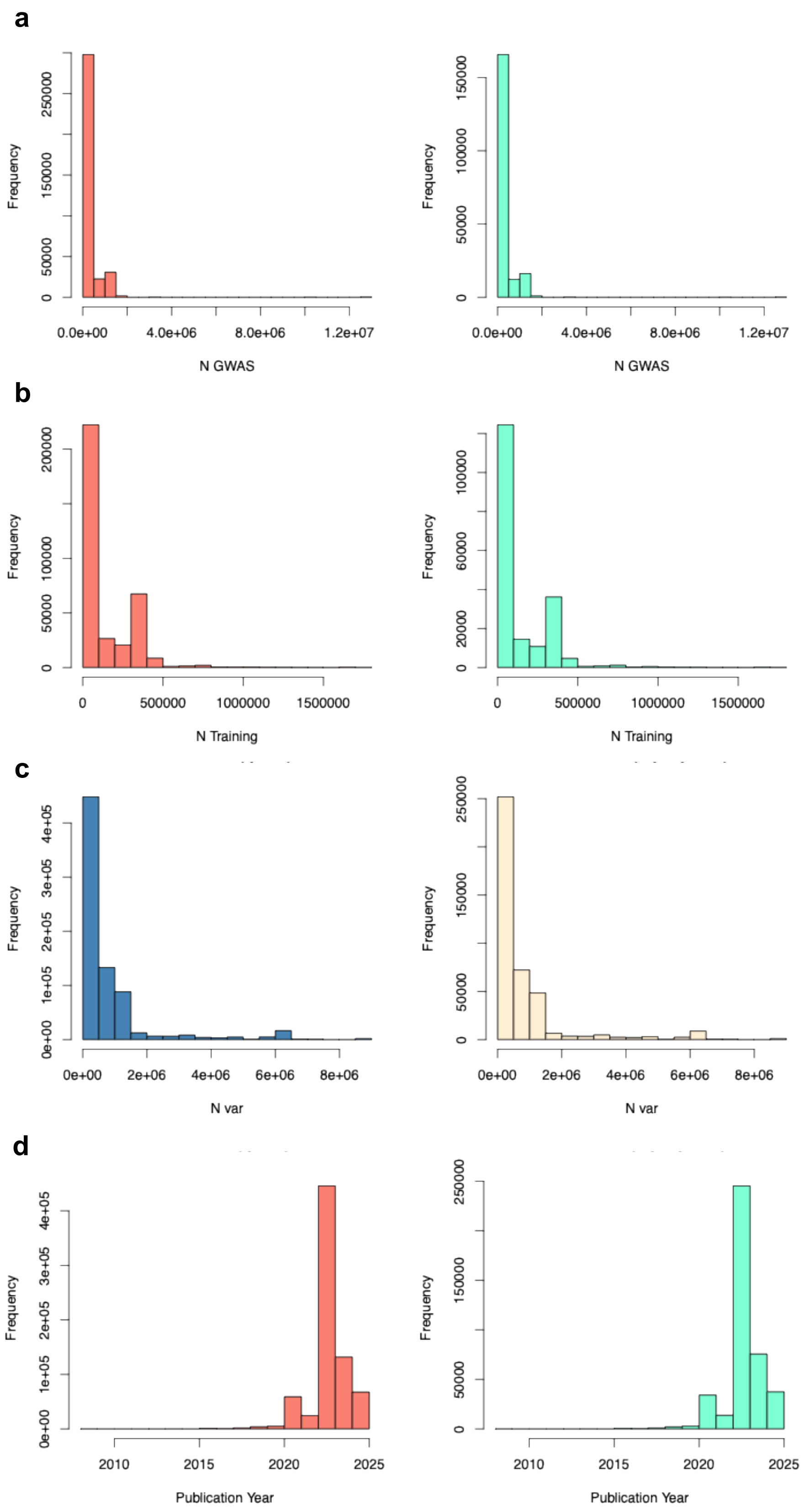
a
Frequency
N GWAS
0.0e+00
4.0e+06
8.0e+06
1.2e+07
b
Frequency
N Training
0
500000
1000000
1500000
c
Frequency
N var
0e+00
2e+06
4e+06
6e+06
8e+06
d
Frequency
Publication Year
2010
2015
2020
2025

**Supplementary Figure 7. Significant (FDR<0.1) features that were log transformed in hierarchical linear mixed models,** as applied to continuous (left) or tresholded unit (right). Such features include **a,** GWAS sample size, **b,** sample size in training cohorts, **c,** number of variants used in the score calculation, **d,** publication year of the scores. Note: more significant associations were found overall for continuous than thresholded models, reflecting differences in counts on y-axes.
